# ChatGPT Assisting Diagnosis of Neuro-ophthalmology Diseases Based on Case Reports

**DOI:** 10.1101/2023.09.13.23295508

**Authors:** Yeganeh Madadi, Mohammad Delsoz, Priscilla A. Lao, Joseph W. Fong, TJ Hollingsworth, Malik Y. Kahook, Siamak Yousefi

## Abstract

**Purpose:** To evaluate the efficiency of large language models (LLMs) including ChatGPT to assist in diagnosing neuro-ophthalmic diseases based on case reports.

**Design:** Prospective study

**Subjects or Participants:** We selected 22 different case reports of neuro-ophthalmic diseases from a publicly available online database. These cases included a wide range of chronic and acute diseases that are commonly seen by neuro-ophthalmic sub-specialists.

**Methods:** We inserted the text from each case as a new prompt into both ChatGPT v3.5 and ChatGPT Plus v4.0 and asked for the most probable diagnosis. We then presented the exact information to two neuro-ophthalmologists and recorded their diagnoses followed by comparison to responses from both versions of ChatGPT.

**Main Outcome Measures:** Diagnostic accuracy in terms of number of correctly diagnosed cases among diagnoses.

**Results:** ChatGPT v3.5, ChatGPT Plus v4.0, and the two neuro-ophthalmologists were correct in 13 (59%), 18 (82%), 19 (86%), and 19 (86%) out of 22 cases, respectively. The agreement between the various diagnostic sources were as follows: ChatGPT v3.5 and ChatGPT Plus v4.0, 13 (59%); ChatGPT v3.5 and the first neuro-ophthalmologist, 12 (55%); ChatGPT v3.5 and the second neuro-ophthalmologist, 12 (55%); ChatGPT Plus v4.0 and the first neuro-ophthalmologist, 17 (77%); ChatGPT Plus v4.0 and the second neuro-ophthalmologist, 16 (73%); and first and second neuro-ophthalmologists 17 (17%).

**Conclusions:** The accuracy of ChatGPT v3.5 and ChatGPT Plus v4.0 in diagnosing patients with neuro-ophthalmic diseases was 59% and 82%, respectively. With further development, ChatGPT Plus v4.0 may have potential to be used in clinical care settings to assist clinicians in providing quick, accurate diagnoses of patients in neuro-ophthalmology. The applicability of using LLMs like ChatGPT in clinical settings that lack access to subspeciality trained neuro-ophthalmologists deserves further research.

**Summary Highlights:**

- The goal of this study was to explore the capabilities of ChatGPT for the diagnoses of different neuro-ophthalmic diseases using specific case examples.
- There was general agreement between ChatGPT Plus v4.0 and two neuro-ophthalmologists in final diagnoses.
- ChatGPT was more general while neuro-ophthalmologists were more methodical and specific when listing diagnoses.

## INTRODUCTION

Neuro-ophthalmology is a subspecialty that bridges the fields of neurology and ophthalmology.^1^ This specialized discipline focuses primarily on conditions affecting visual pathways, visual processing, and eye movements. Practitioners in this subspecialty have completed residency in neurology or ophthalmology and then an additional year of fellowship training in neuro-ophthalmology. Neuro-ophthalmology is a cognitively intense field, requiring a detailed history and examination and synthesis of neuro-imaging results and lab work, to diagnose often vision- and life-threatening conditions. Neuro-ophthalmologists often are a “specialist’s specialist,” where many patients are referred by ophthalmologists or neurologists. In a prospective study, nearly half of all referrals to neuro-ophthalmology carried an incorrect diagnosis, and a quarter of these misdiagnosed patients suffered harm.^2^

To add further insult to injury, neuro-ophthalmologists are in short supply, deemed a “human resource crisis” since at least 2008.^3^ As of 2022, only 8 states in the United States have adequate coverage and the median wait time to see a neuro-ophthalmologist is 6 weeks.^4^

In recent years, artificial intelligence (AI) and machine learning (ML) technologies have provided promising solutions in various healthcare domains.^5^ Several studies have shown the efficacy of AI models in ophthalmology.^6-8^ More recently, emerging large language models (LLMs) including ChatGPT (OpenAI, LLC, California, USA), have garnered a great deal of attention for their ability to comprehend and generate human-like and fluent text responses to queries.^9^ The commercially available ChatGPT Plus 4.0 has set a new standard for text generation with unparalleled fluency and human-likeness that stretches these limits even further than its predecessor, ChatGPT 3.5. It is worth mentioning that ChatGPT 3.5 was initially trained based on an impressive corpus of over 400 billion words gathered from the web, including books, articles, and diverse digital content.

ChatGPT (accessible at https://chat.openai.com), leverages the architecture of the ChatGPT Plus 4.0 and responds to textual input with high precision and nuance due to improved finetuning. The ChatGPT iteration of ChatGPT Plus 4.0 is endowed with an expanded toolkit of natural language processing (NLP) features, ranging from more accurate translation to sophisticated text summarization, demonstrating OpenAI’s commitment to advancing the state of conversational AI.

Efforts now have been made to leverage ChatGPT’s capabilities in specialized domains, such as medical diagnostics, in addition to general text interactions.^10^ However, there are handful research studies discussing applications of ChatGPT in Ophthalmology^11, 12, 13, 14^ and the efficacy and reliability of ChatGPT in the specialized field of ophthalmology including neuro-ophthalmology have not been adequately investigated. This paper aims to evaluate ChatGPT’s diagnostic capabilities in the field of neuro-ophthalmology. We intend to investigate the model’s capabilities and limitations by comparing responses to detailed case descriptions from several patients with various neuro-ophthalmic conditions.

## METHODS

### Case Collection

We utilized cases from the Department of Ophthalmology and Visual Sciences at the University of Iowa’s publicly accessible database (https://webeye.ophth.uiowa.edu/eyeforum/cases.html). We selected 22 cases with various common and uncommon neuro-ophthalmic diseases including “Acute Demyelinating Encephalomyelitis (ADEM) with Associated Optic Neuritis”, “Optic Nerve Drusen”, “Optic Nerve Hypoplasia”, “Optic Neuritis”, “Optic Nerve Hypoplasia”, “Cranial Nerve IV (Trochlear Nerve) Palsy”, “Dorsal Midbrain Syndrome (Parinaud’s Syndrome)”, “Traumatic Optic Neuropathy”, “Ethambutol Toxicity and Optic Neuropathy”, “Posterior Ischemic Optic Neuropathy”, “Recurrent Neuroretinitis”, “Horner’s Syndrome due to Cluster Headache”, “Horner Syndrome due to Ipsilateral Internal Carotid Artery Dissection”, “Idiopathic Intracranial Hypertension (Pseudotumor Cerebri)”, “Idiopathic Orbital Myositis”, “Leber Hereditary Optic Neuropathy”, “Miller Fisher Syndrome”, “Myasthenia Gravis”, “Neurofibromatosis Type 1—Optic Nerve Glioma”, “Thyroid Eye Disease (Graves’ ophthalmopathy)”, “Visual Snow Syndrome”, and “Unilateral Optic Nerve Hypoplasia”. Descriptions of each case included patient demographics, history of the presenting illness, chief complaint, relevant medical or ocular history, and examination findings.

Our local institutional review board (IRB) office advised that no IRB approval is required for this study because we utilized a publicly available dataset containing no patient information and the study adhered to the Helsinki declaration’s principles.

### ChatGPT for Diagnosis

We feed each case description into the ChatGPT (versions 3.5 and 4.0) and asked if the model could provide a diagnosis. We specifically asked: “What is the most probable diagnosis?” (Figures 1 and 2). Then we evaluated the accuracy of ChatGPTs in making a correct diagnosis. As ChatGPT may learn from previous interactions once a single chatBox is used, we recorded all responses based on our first inquiry of diagnosis and used a new chatBox in any case that we asked same questions more than once.

**Figure 1.**
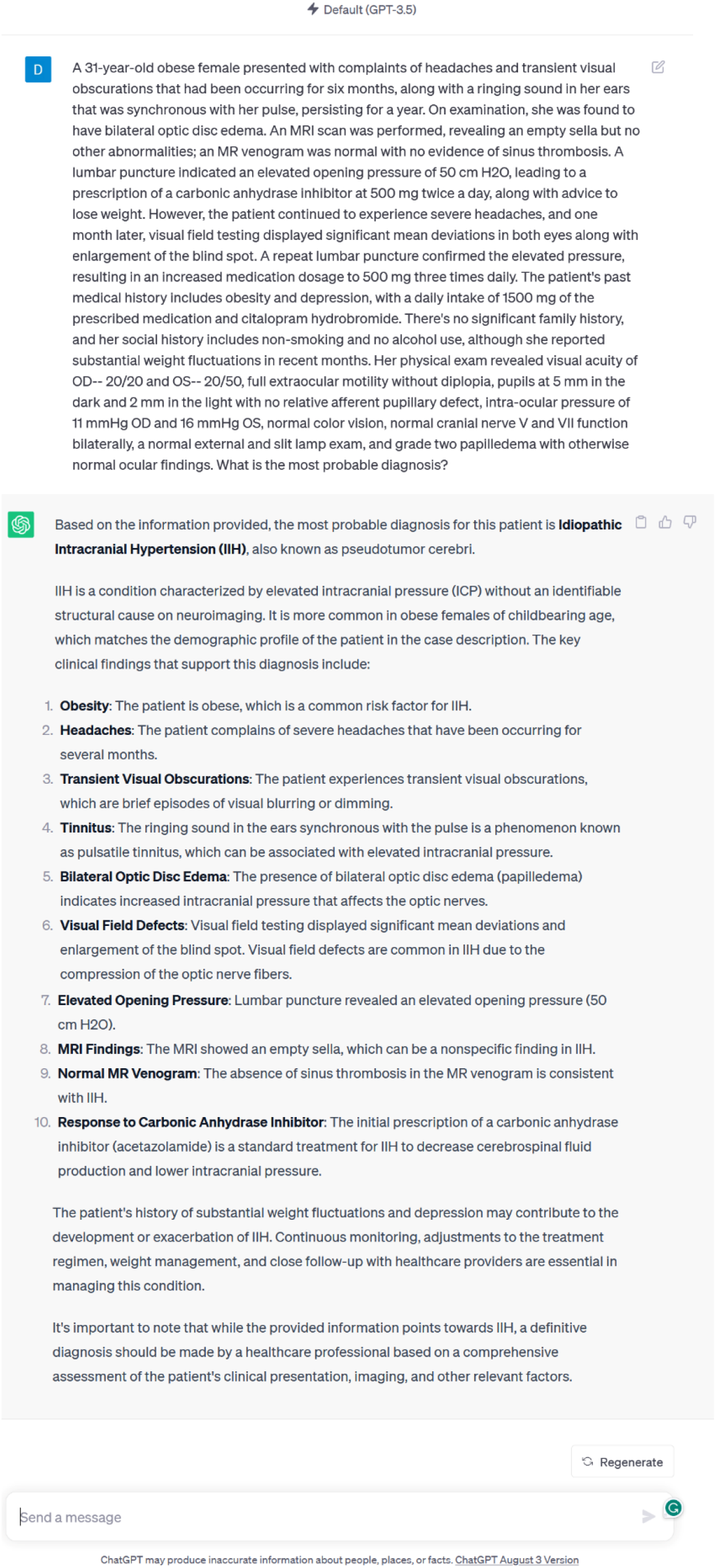
A sample case description of a patient with idiopathic intracranial hypertension (pseudotumor cerebri) input into the ChatGPT 3.5 model and the corresponding response.

**Figure 2.**
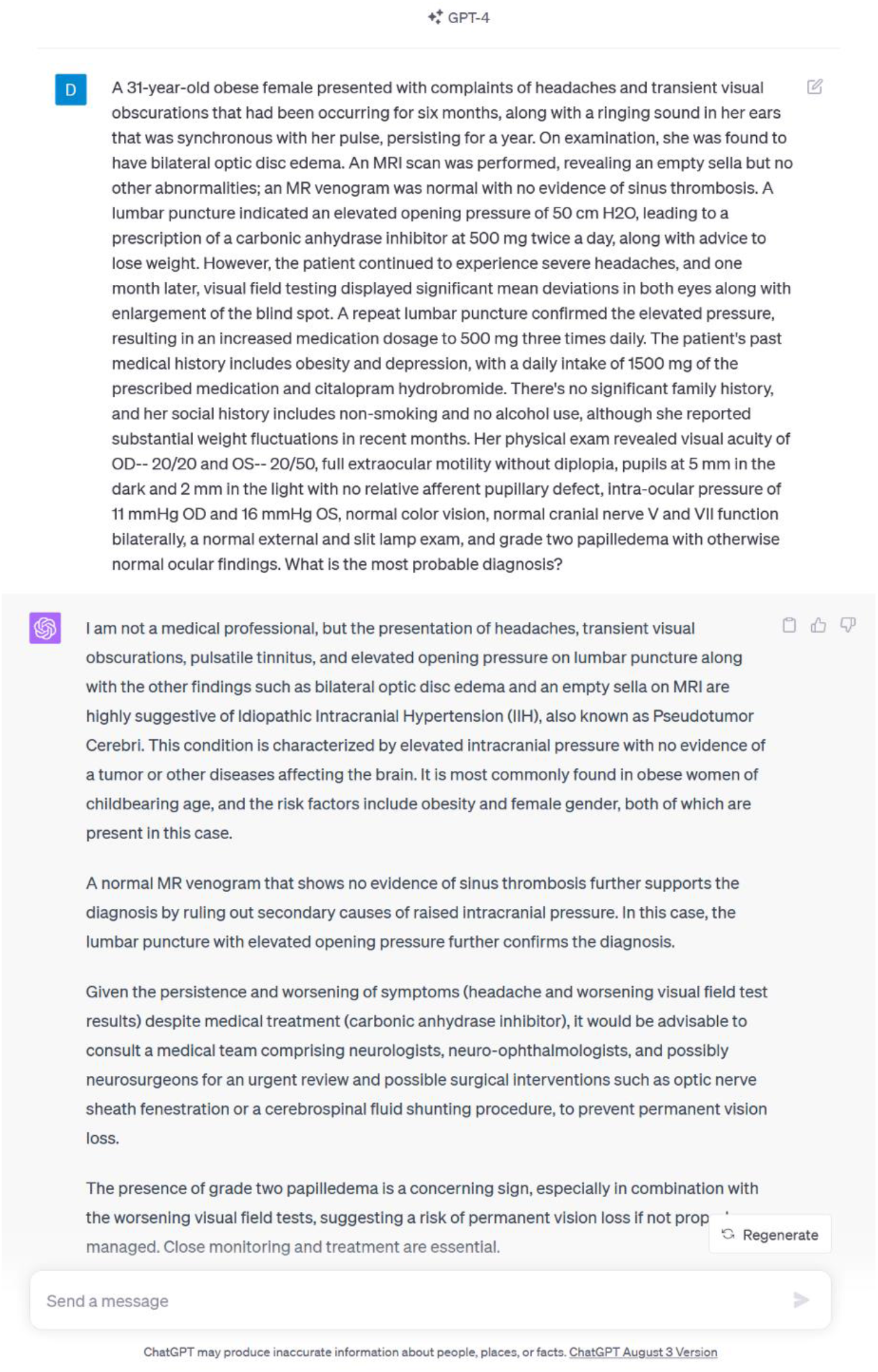
A sample case description of a patient with idiopathic intracranial hypertension (pseudotumor cerebri)) input into the ChatGPT Plus 4.0 model and the corresponding response.

Additionally, we provided the same 22 cases to two neuro-ophthalmologists in our institute in a masked manner and asked them to make provisional diagnosis. Neuro-ophthalmologists were not allowed to use ChatGPT or other similar tools for making diagnosis. We then computed the frequency of correct diagnoses of ChatGPTs and the two neuro-ophthalmologists and compared the accuracies. We also evaluated the agreement between ChatGPTs and the two neuro-ophthalmologists.

## RESULTS

ChatGPT 3.5 and ChatGPT Plus 4.0 made the correct diagnosis in 13 (59%) and 18 (82%) out of 22 cases, respectively, while two neuro-ophthalmologists both correctly diagnosed 19 (86%) out of 22 cases. Table 1 represents the details of diagnoses which are provided by ChatGPT 3.5, ChatGPT Plus 4.0 and the two neuro-ophthalmologists.

**Table 1.**
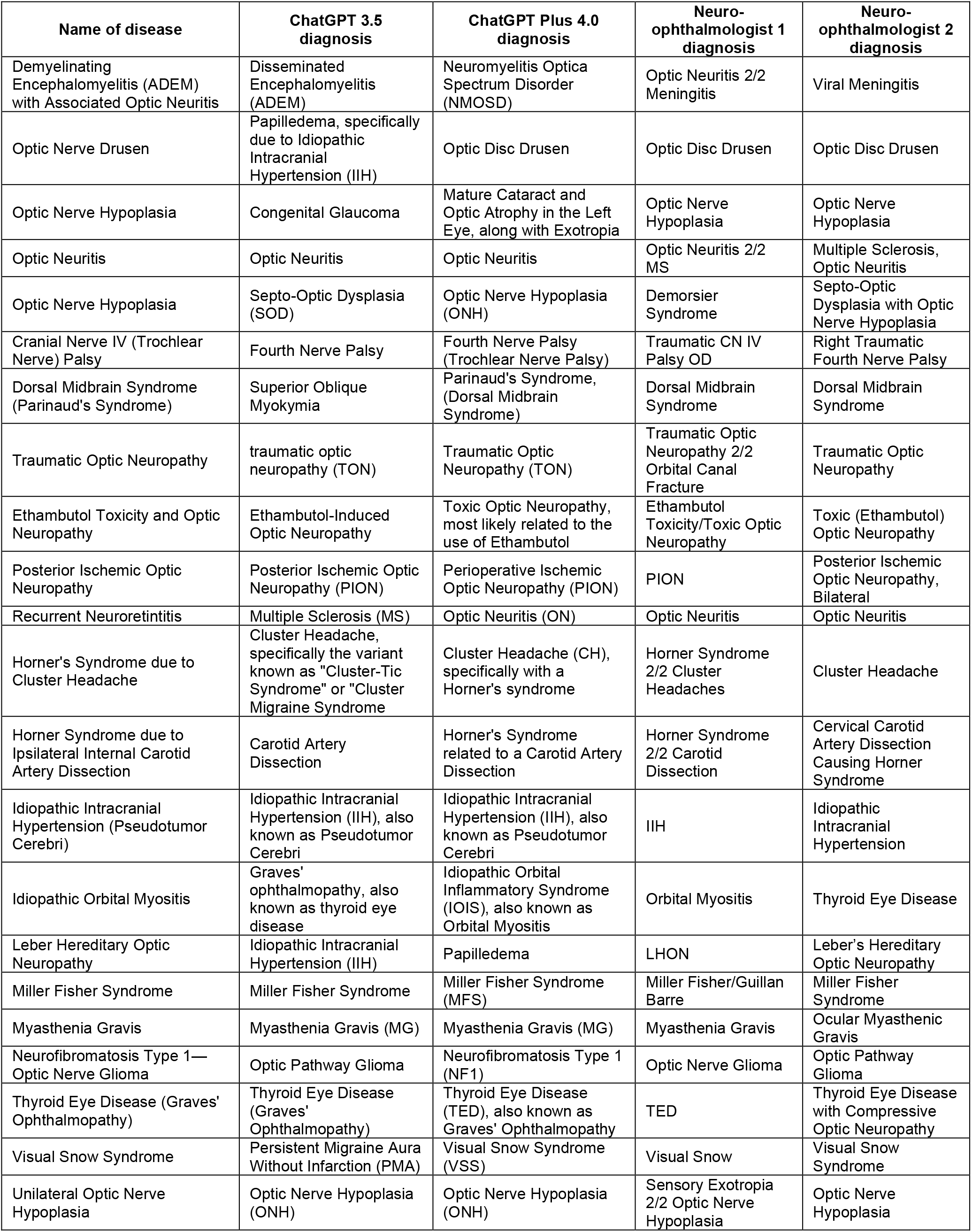
Diagnoses provided by ChatGPT 3.5, ChatGPT Plus 4.0, and two neuro-ophthalmologists.

The agreement between the various diagnostic sources were as follows: ChatGPT v3.5 and ChatGPT Plus v4.0, 13 (59%); ChatGPT v3.5 and the first neuro-ophthalmologist, 12 (55%); ChatGPT v3.5 and the second neuro-ophthalmologist, 12 (55%); ChatGPT Plus v4.0 and the first neuro-ophthalmologist, 17 (77%); ChatGPT Plus v4.0 and the second neuro-ophthalmologist, 16 (73%); and first and second neuro-ophthalmologists 17 (17%).

## DISCUSSION

We prospectively investigated the diagnostic capability of ChatGPT in neuro-ophthalmic diseases based on 22 cases collected from the University of Iowa online database. ChatGPT 3.5, ChatGPT Plus 4.0, and both neuro-ophthalmologists diagnosed 13, 18, 19, and 19 cases correctly out of 22 cases, respectively. Despite the complexity of neuro-ophthalmic cases, the diagnostic capability of ChatGPT Plus 4.0 was close to neuro-ophthalmologists. Under active training, it is expected that newer versions of ChatGPT may become more accurate in the future.

While the agreements between ChatGPT v3.5 and neuro-ophthalmologists were relatively low (55% - 59%), ChatGPT Plus v4.0 had a reasonable agreement with neuro-ophthalmologists (73% - 77%). The agreement between the neuro-ophthalmologists was 77%, on par with ChatGPT Plus 4.0.

Understanding capabilities and limitations of ChatGPT will help to effectively utilize this technology in neuro-ophthalmology research and clinical practice.

A recent study assessed the performance of ChatGPT in answering practice questions in neuro-ophthalmology and observed that ChatGPT was correct on 3 out of 7 responses (43%) of the text-based questions.^15^ Another study investigated ChatGPT 3.5 and ChatGPT Plus 4.0 capabilities in responding to various ophthalmological questions including neuro-ophthalmic questions based on the online database of OphthoQuestions and the American Academy of Ophthalmology’s Basic Clinical Science Course (BCSC). They identified that the accuracy of ChatGPT 3.5 was 10% and 32% and the accuracy of ChatGPT Plus 4.0 was 40% and 48% based on the OphthoQuestions and BCSC neuro-ophthalmic questions, respectively.^11^ Their reported accuracy in ChatGPT capability in responding to neuro-ophthalmic questions is significantly lower than what we observed in neuro-ophthalmic diagnosis, however, the nature of these two studies are different, making it challenging to directly comparing the findings of these two studies.

Advancement of LMM models including ChatGPT may benefit ophthalmology in general and neuro-ophthalmology in particular by making more objective and quick diagnosis based on readily accessible models. Neuro-ophthalmic issues result from a complex interaction between the nervous system and visual apparatus, necessitating a comprehensive evaluation to identify subtle distinctions.^16^

Another significant advantage of ChatGPT and comparable LMM models is that they can actively learn based on their reinforcement learning capability, thereby correcting their previous errors, enhancing their performance over time, and becoming more precise. In addition, LMM models require minimal human monitoring and supervision for active training, which is advantageous in comparison to supervised learning models that require intensive human oversight.

While we used patients with various phenotypes of neuro-ophthalmic conditions, our study does have some limitations. First, we used cases from an online database to investigate ChatGPT capabilities raising concerns that ChatGPT may have previously been exposed to this database. To resolve this issue, we examined the years in which these cases were added to the database and found that the vast majority were added before September 2021, when the most recent ChatGPT training concluded. Versions 3.5 and 4.0 of ChatGPTs contained errors in 9 and 4 instances, respectively, alleviating concerns that this database has been accessed previously. Second, ChatGPT has been evaluated based on 22 cases. Consequently, other studies may evaluate ChatGPT on the basis of a larger number of cases to confirm our findings. Another limitation of LLMs like ChatGPT is lack of image interpretation that limits their applicability in more complex cases with information from various imaging modalities. Additionally, while these were real world cases, they were published primarily for teaching purposes. Several of these cases exhibited classic presentations of diseases with “buzzword” descriptions with definitive diagnoses. Therefore, this subset of cases may not fully reflect the actuality of neuro-ophthalmology practice with borderline findings and confounding factors in many instances. Conversely, some cases exhibited atypical presentations of the actual diagnosis. This is where the art of medicine comes into play and requires a human practitioner to weigh these factors appropriately.

In summary, various AI models have shown great potential in ophthalmology. Emerging ChatGPT has been recently applied to ophthalmic education as well as diagnosis and has been shown to be capable of conversing with patients as well as physicians and respond to text inquiries with fluent and reasonable accuracy. These models may also be utilized in a variety of healthcare settings, including primary care offices or emergency services to aid in patient triaging, with the caveat that not all patient information is available in such settings. These models can also be useful in tertiary ophthalmology care, to provide initial assessments. Once capable of interpreting images in the future, LMMs including ChatGPT may provide more objective, rapid, and possibly accurate diagnosis based on both textual and imaging input.

## CONCLUSION

The introduction of LMMs, such as ChatGPT, represents a potentially revolutionary step forward augmenting clinical diagnostic, independent of the availability of subspeciality trained clinicians. This is especially interesting in fields such as neuro-ophthalmology, which is characterized by an abundance of complicated cases and the absence of a conclusive diagnostic testing. ChatGPT Plus v4.0 appears capable of diagnosing complex neuro-ophthalmic cases, when provided with structured data in case report format, with a level of accuracy that is comparable to that of experienced neuro-ophthalmologists. Re-assessment of the performance of ChatGPT, and other LLMs, in similar testing conditions but with multimodal capabilities would be a natural next research step when practical and pending availability of such algorithms. LLMs could potentially bridge the gap between AI and human competence. Future research should explore how technological innovations, such as enhanced LLMs, can augment medical decision making and assist clinicians towards enhancing patient outcomes.

## Data Availability

All data produced are available online at the University of Iowa's publicly accessible database (https://webeye.ophth.uiowa.edu/eyeforum/cases.html).

## Acknowledgement

This work was supported by NIH Grants R01EY033005 (SY), R21EY031725 (SY), and grants from Research to Prevent Blindness (RPB), New York (SY). The funders had no role in study design, data collection and analysis, decision to publish, or preparation of the manuscript.

## REFERENCES

1. Levin LA, Nilsson SF, Ver Hoeve J, Wu S, Kaufman PL, Alm A. Adler’s Physiology of the Eye E-Book: Expert Consult-Online and Print. Elsevier Health Sciences; 2011.

2. Stunkel L, Sharma RA, Mackay DD, et al. Patient harm due to diagnostic error of neuro-ophthalmologic conditions. Ophthalmology. 2021;128(9):1356–1362.

3. Frohman LP. The human resource crisis in neuro-ophthalmology. Journal of Neuro-Ophthalmology. 2008;28(3):231–234.

4. DeBusk A, Subramanian PS, Scannell Bryan M, et al. Mismatch in supply and demand for neuro-ophthalmic care. Journal of Neuro-Ophthalmology. 2022;42(1):62–67.

5. Esteva A, Robicquet A, Ramsundar B, et al. A guide to deep learning in healthcare. Nature medicine. 2019;25(1):24–29.

6. Kapoor R, Walters SP, Al-Aswad LA. The current state of artificial intelligence in ophthalmology. Survey of ophthalmology. 2019;64(2):233–240.

7. Li J-PO, Liu H, Ting DS, et al. Digital technology, tele-medicine and artificial intelligence in ophthalmology: A global perspective. Progress in retinal and eye research. 2021;82:100900.

8. Li Z, Wang L, Wu X, et al. Artificial intelligence in ophthalmology: The path to the real-world clinic. Cell Reports Medicine. 2023;

9. Brown T, Mann B, Ryder N, et al. Language models are few-shot learners. Advances in neural information processing systems. 2020;33:1877–1901.

10. Obermeyer Z, Emanuel EJ. Predicting the future—big data, machine learning, and clinical medicine. The New England journal of medicine. 2016;375(13):1216.

11. Antaki F, Touma S, Milad D, El-Khoury J, Duval R. Evaluating the Performance of ChatGPT in Ophthalmology: An Analysis of Its Successes and Shortcomings. Ophthalmol Sci. Dec 2023;3(4):100324. doi:10.1016/j.xops.2023.100324

12. Singh S, Djalilian A, Ali MJ. ChatGPT and Ophthalmology: Exploring Its Potential with Discharge Summaries and Operative Notes. Semin Ophthalmol. Jul 2023;38(5):503–507. doi:10.1080/08820538.2023.2209166

13. Cai LZ, Shaheen A, Jin A, et al. Performance of Generative Large Language Models on Ophthalmology Board-Style Questions. Am J Ophthalmol. Jun 18 2023;254:141–149. doi:10.1016/j.ajo.2023.05.024

14. Delsoz M, Madadi Y, Munir WM, et al. Performance of ChatGPT in Diagnosis of Corneal Eye Diseases. medRxiv. 2023:2023.08.25.23294635. doi:10.1101/2023.08.25.23294635

15. Mihalache A, Popovic MM, Muni RH. Performance of an Artificial Intelligence Chatbot in Ophthalmic Knowledge Assessment. JAMA Ophthalmol. Jun 1 2023;141(6):589–597. doi:10.1001/jamaophthalmol.2023.1144

16. Liu GT, Volpe NJ, Galetta SL. Neuro-ophthalmology e-book: Diagnosis and management. Elsevier Health Sciences; 2010.

